# Deep learning ensemble for abdominal aortic calcification scoring from lumbar spine X-ray and DXA images

**DOI:** 10.1101/2025.04.23.25326192

**Authors:** Antti Voss, Sanna Suoranta, Tomi Nissinen, Ossi Hurskainen, Amro Masarwah, Reijo Sund, Jussi Tohka, Sami P Väänänen

## Abstract

Abdominal aortic calcification (AAC) is an independent predictor of cardiovascular diseases (CVDs). AAC is typically detected as an incidental finding in spine scans. Early detection of AAC through opportunistic screening using any available imaging modalities could help identify individuals with a higher risk of developing clinical CVDs. However, AAC is not routinely assessed in clinics, and manual scoring from projection images is time-consuming and prone to inter-rater variability. Also, automated AAC scoring methods exist, but earlier methods have not accounted for the inherent variability in AAC scoring and were developed for a single imaging modality at a time.

We propose an automated method for quantifying AAC from lumbar spine X-ray and Dual-energy X-ray Absorptiometry (DXA) images using an ensemble of convolutional neural network models that predicts a distribution of probable AAC scores. We treat AAC score as a normally distributed random variable to account for the variability of manual scoring. The mean and variance of the assumed normal AAC distributions are estimated based on manual annotations, and the models in the ensemble are trained by simulating AAC scores from these distributions. Our proposed ensemble approach successfully extracted AAC scores from both X-ray and DXA images with predicted score distributions demonstrating strong agreement with manual annotations, as evidenced by concordance correlation coefficients of 0.930 for X-ray and 0.912 for DXA. The prediction error between the average estimates of our approach and the average manual annotations was lower than the errors reported previously, highlighting the benefit of incorporating uncertainty in AAC scoring.

## 1. Introduction

Globally, cardiovascular diseases (CVDs) are responsible for approximately 30% of all deaths, claiming nearly 18 million lives each year *[1]*. The most prevalent form of CVDs is atherosclerosis, causing arteries to narrow and harden from plaque accumulation. Arterial calcification, particularly abdominal aortic calcification (AAC) is a manifestation of atherosclerosis and an independent prognostic indicator and risk factor for future CVD events and mortality *[2, 3, 4, 5]*. The abdominal aorta is also one of the initial sites where calcification is seen, and its presence is a sign of generalized atherosclerosis elsewhere in the arterial system *[6, 7]*. Atherosclerosis and calcification are long-lasting dynamic processes, and it usually takes decades to develop CVD events, such as heart attack or stroke. Thus, early detection of AAC through opportunistic screening can help identify people with increased CVD risk and start preventive measures on time.

Medical imaging is one of the few options to detect AAC, and typically AAC is discovered as an incidental finding in a scan that examines the spine or abdomen region. AAC can be detected using various imaging modalities such as Computed Tomography (CT) *[8, 9, 10]*, conventional X-ray *[11, 9, 12]*, and Dual-energy X-ray Absorptiometry (DXA) *[8, 13, 14, 15, 9, 16, 17]*. CT is the most accurate technique to quantify AAC, and several automated methods based on it have been developed, e.g., *[18, 19, 20, 21]*. However, CT imaging is expensive and causes a high radiation burden. Lumbar spine lateral X-rays and DXA scans can also reveal AAC, using a lower radiation dose compared to CT scans. Although X-ray and DXA cannot represent the calcification volumetrically, the lateral projection images can still quantify AAC with a good correlation to CT *[8, 9]*. To achieve optimal opportunistic screening coverage, it would be essential to automatically detect AAC from X-ray and DXA images. In this paper, we propose a method to detect AAC from these imaging modalities.

The most widely used method for assessing the AAC from lateral projection images is the semi-quantitative 24-point scoring method developed by Kauppila et al. *[11]*. In this method, the calcification in the aorta is measured visually at lumbar vertebra levels L1 to L4, where each vertebra level receives a score from 0 to 6. The total AAC score, ranging from 0 to 24 (AAC24 score), is the sum of these individual scores. Manual scoring of AAC from X-ray and DXA images is both labor-intensive and subjective *[11, 16]*. As a result, currently, AAC is not routinely screened or reported by clinicians from X-ray or DXA images.

Previous approaches for automating AAC scoring in X-ray *[22, 23]* and DXA *[24, 25, 26, 27, 28, 29, 30, 31, 32]* images have focused on machine learning and deep neural network methods. In most of them, the AAC scoring task has been formulated either as a segmentation problem or as a regression task to directly estimate the cumulative AAC24 score. More recently, methods that estimate AAC separately at vertebra levels L1 to L4 have been proposed *[30, 31, 28]*. However, in all these studies, the manually scored and automatically estimated AAC scores have been treated as fixed definite values, despite only moderate to good inter-rater agreement between two individual scorers *[11, 16]*. In addition, earlier methods have been developed for a single imaging modality at a time. Since the visibility of AAC differs in images of different modalities, AAC scores predicted from different modalities are not directly comparable.

To overcome the abovementioned limitations in automated AAC scoring methods, we propose a sampling-based training approach combined with an ensemble of convolutional neural network (CNN) models to estimate AAC from both X-ray and DXA images. The inherent variability in the annotated AAC scores is considered by treating the scores as normally distributed random variables instead of deterministic point values. The variability and uncertainty in AAC scores are captured in the ensemble of CNN models, which predicts a distribution of scores that approximates AAC at vertebra levels L1 to L4.

## 2. Materials and Methods

### 2.1 Training and external test set images

We used two separate imaging data sets collected from two different hospitals, one for model selection, training, and internal testing, and the other for external testing of the model ensemble performance. For training, 2172 lateral lumbar spine X-ray and DXA image pairs (from 2172 patients) were obtained from Joensuu central hospital (Wellbeing services county of North Karelia, Finland), and for external testing a smaller subset of 235 similar image pairs (from 235 patients) from a large set of images were collected from Kuopio University Hospital (Wellbeing services county of North Savo, Finland). The details of these data sets are illustrated in Fig. 1. Images for the study were collected according to a research permission granted by the Finnish Institute for Health and Welfare (THL/2247/5.05.00/2019) and ethical permission granted by Research Ethics Committee of Hospital District of Northern Savo (1298/13.02.00/2019).

**Figure 1:**
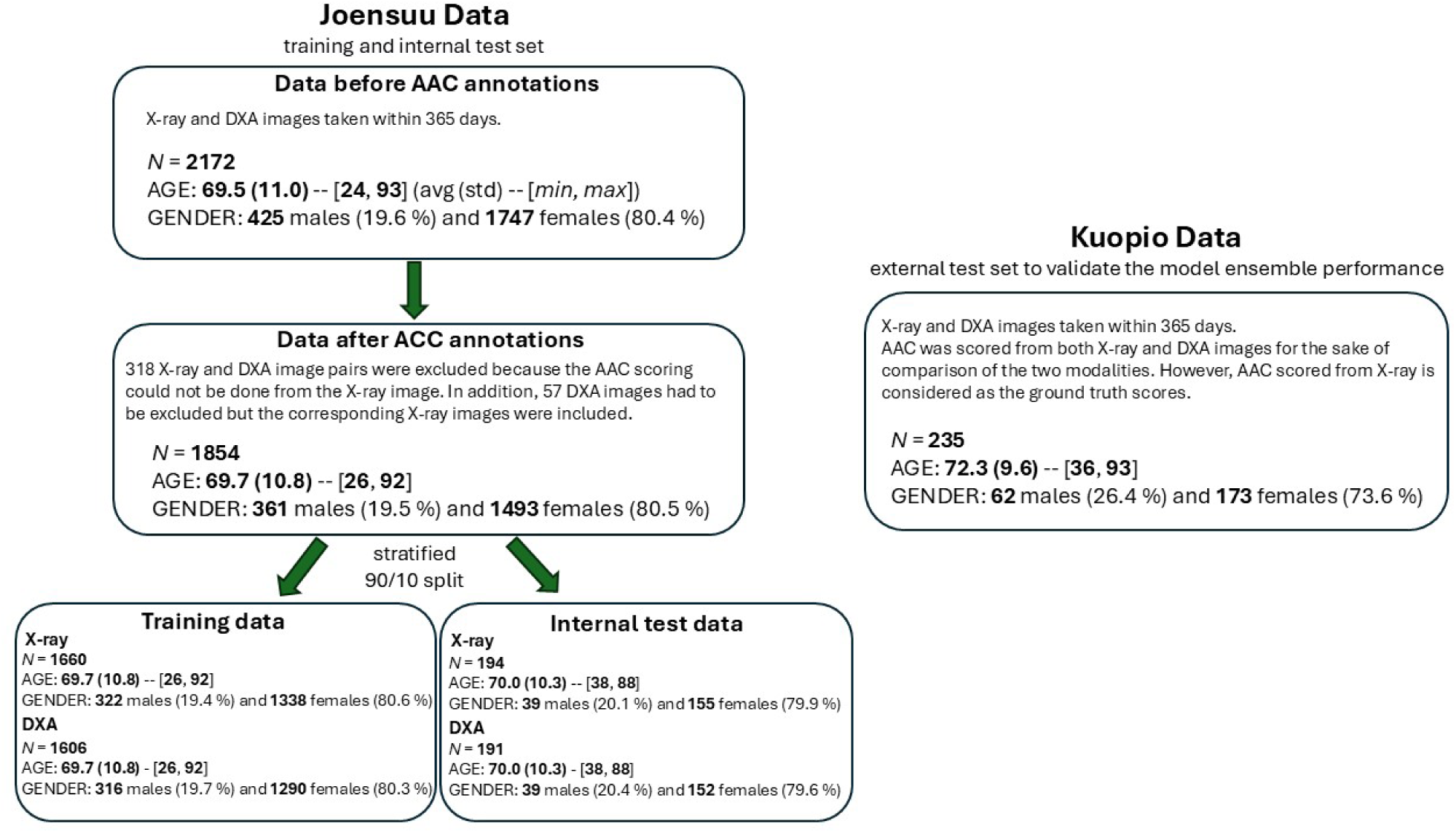
Details of the two separate imaging data sets used in this study.

X-ray images were acquired with scanners from different manufacturers (Philips, Fujifilm Corporation, Kodak, Carestream Health, Siemens, Agfa and Samsung) and energies ranging from 78 to 102 kV. X-ray images were scaled to 0.14 mm pixel size. DXA images were taken with GE Lunar iDXA scanners, and their original pixel size 0.60 x 0.25 mm was scaled to 0.60 x 0.60 mm. One DXA scan included three images: two measured with low (35 kV) and high (70 kV) energies and a bone mineral density (BMD) image, which is computed based on the low and high energy images *[33]*.

### 2.2 AAC annotations and datasets

To assess the extent of AAC, we used a 24-point semi-quantitative scoring scale developed by Kauppila et. al *[11]* for lumbar spine X-ray images. The AAC24-score evaluates the calcification in the anterior and posterior walls of the abdominal aorta at the lumbar vertebra L1 to L4 levels. Within each of these 8 segments, the scoring of aortic calcification is based on comparing the total height of the calcification in the aortic wall to the height of the adjacent lumbar vertebra. Each segment is scored from 0 to 3. The segment receives a score of 0 if there is no calcification. If the calcification covers less than 1/3 of vertebral height, the segment is scored as 1. The segment is scored as 2 if the total height of calcification is between 1/3 and 2/3 of vertebral height. Finally, the segment is scored as 3, if the calcification height exceeds 2/3 of the vertebral height. This results in an overall score of 0 to 24. AAC score adjacent to each vertebra is referred to as *AAC6*, and the combined total score as *AAC24*.

The X-ray-DXA image pairs included in the study had their scan dates within 365 days from each other. During that period, AAC can be expected to change very little because it generally develops at a slow pace *[34, 35]*. Five annotators (two radiologists, two researchers, and a medical physicist) scored AAC from the training data set (Joensuu set) X-rays. The AAC scored by non-radiologists was found to have similar variability than the scores made by radiologists. The AAC scoring of the external test set (Kuopio set) was done by three radiologists, each providing two annotated scores for each image on separate occasions. In contrast to the Joensuu set, AAC was scored for the Kuopio set from both X-ray and DXA images.

From the Joensuu set, 1854 X-ray images out of 2172 were included for training, model selection and internal testing after AAC scoring. An image was included if 3 out of 5 annotators had scored the AAC from the image. AAC was not annotated if an annotator interpreted that the image had artifacts that made reading the image impossible or the region where the aorta was expected to be located was not visible. For DXA images, 1797 out of 1854 were available for training, since 57 DXA images had to be excluded due to the exclusion criteria mentioned above. The division to training and internal test sets was done in a stratified manner, meaning that the distributions of the mean AAC24 were similar in both sets. 90 percent, i.e., 1660 X-ray images were included in the training set and 10 percent, i.e., 194 images in the internal test set. For DXA images, the number of images in the training and internal test sets were 1606 and 191, respectively.

For the external test set, AAC was scored from all 235 X-ray and DXA images. The X-ray-DXA image pairs were selected randomly from a large set of suitable pairs, and before inclusion, each image pair was visually checked to ensure that the aortic region was visible. The model performance was evaluated using the annotated scores from X-ray images, whereas the annotated scores from DXA images were only used to compare the agreement between annotated AAC values scored from X-ray and DXA images.

### 2.3 Estimation of AAC distribution parameters

Fig. 2. shows an example of AAC scoring for one X-ray image from the training set and its DXA image pair. For this X-ray image, we calculated the AAC6 score statistics (mean and variance) from AAC scores made by 5 annotators. Each AAC6 score is assumed as normally distributed, and the four scores are assumed to be independent. For each vertebra level, the mean and variance of the normal distribution are estimated using all AAC annotations for that specific vertebra level. Fig. 2. also shows these individual annotated AAC6 scores, and the approximated normal distributions at the vertebra levels, named as *AAC6 distributions*, together with the total AAC24 score and its distribution, of which the latter is obtained by adding the four vertebra-level normal distributions. The estimation of the mean and variance of AAC6 distributions was done separately for all annotated images.

**Figure 2:**
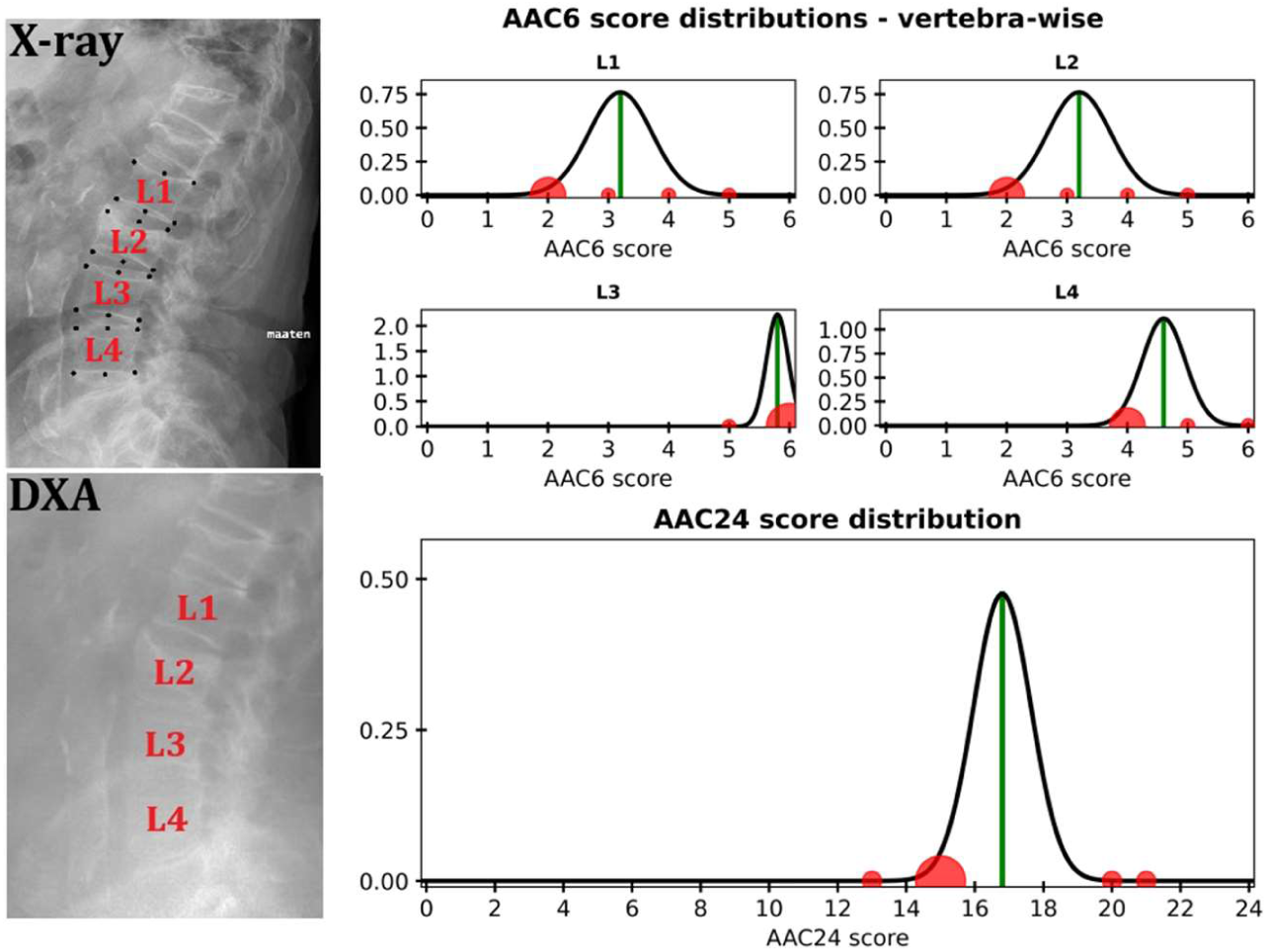
Left: Lateral lumbar spine X-ray radiograph and the corresponding DXA image. Right: The scored AAC values (red dots) are shown for each vertebra level and for the AAC24 score. Green lines illustrate the average AAC6 and AAC24 scores and the black distributions illustrate the approximated AAC6 and AAC24 normal distributions.

### 2.4 Deep learning approach to estimate AAC

Our automated algorithm for estimating the AAC distributions from lateral lumbar spine X-ray or DXA images has two phases (Fig 3.): X-ray/DXA image cropping and AAC scoring from the cropped image. In the first phase, the algorithm locates the region where abdominal aorta is expected to be anterior to vertebrae L1 to L4. The cropping of the aortic area was done by defining a region of interest based on landmark points that were estimated using CNN-based deep-learning models. The details of the landmark estimation and cropping are presented in the Supplementary material. In the second phase, the cropped image is fed to an ensemble of models where each model outputs four AAC6 scores, one for each vertebra level L1 to L4. Since each model in the ensemble is trained with outputs that are sampled from the estimated AAC6 normal distributions, the ensemble of models produces an estimate for each of the four AAC6 distributions. The total AAC24 distribution is obtained by summing the four AAC6 distributions together. And since the ensemble outputs distributions of probable AAC scores, one can also estimate confidence intervals for any point estimates, such as the mean, to assess the reliability of the estimation.

**Figure 3:**
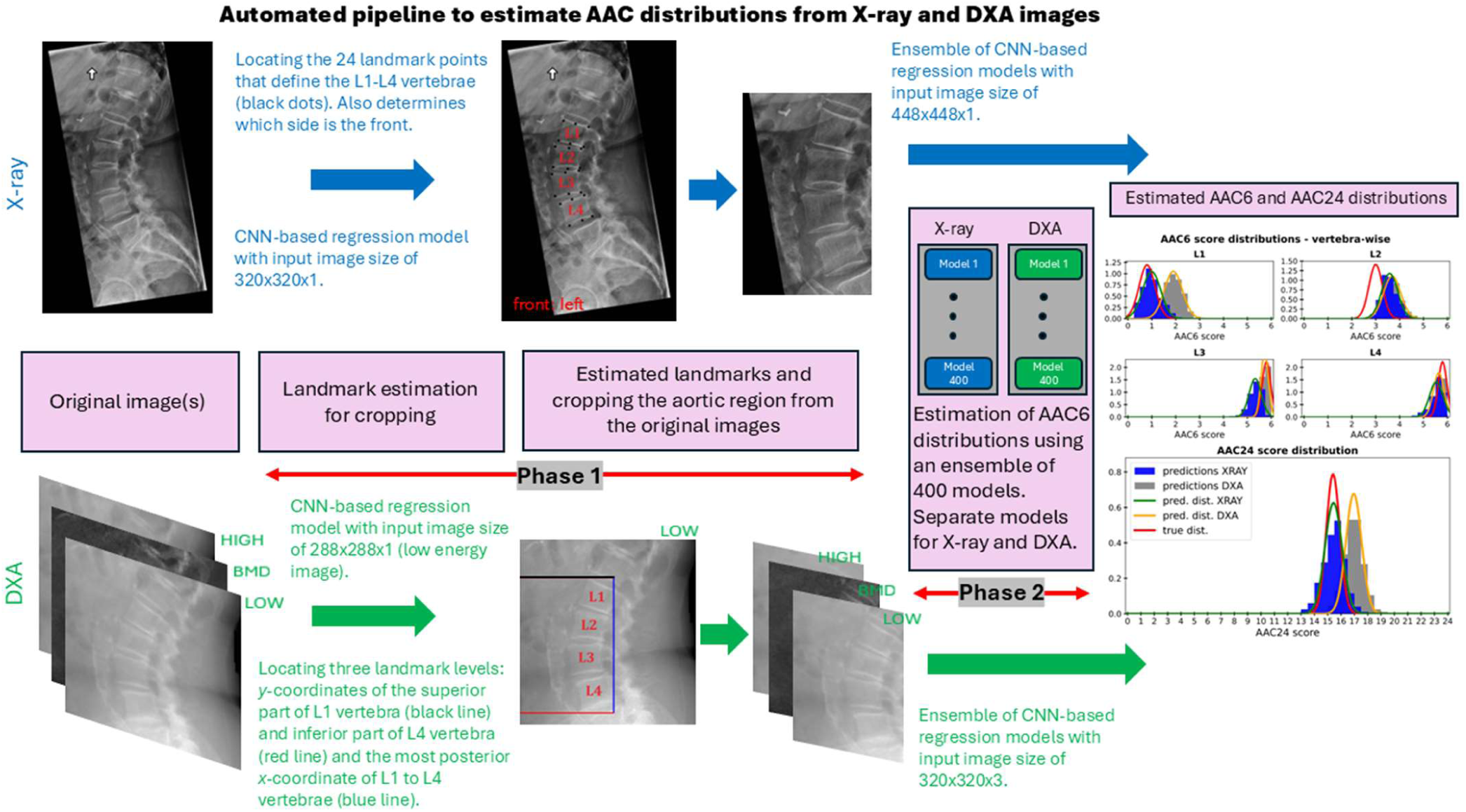
A pipeline to estimate the AAC6 distributions using a CNN-based deep learning model ensemble for X-ray and DXA images.

After the original images were cropped, the process of training the model ensemble to estimate the AAC6 distributions was the same for X-ray and DXA images. First, the training data was divided into 10 folds using stratified sampling (similar mean AAC24 distributions in each fold). For each set of fold divisions (9 folds for training and 1 for validation) we trained 40 models, and repeating this to all possible fold combinations resulted in an ensemble consisting of 400 models.

For each individual model and all its training images, two data augmentation steps were performed. First, an image was augmented using: rotation ([-15°, 15°]), shear ([-5°, 5°]), translation (*x*:[-15%, 15%], *y*:[-10%, 10%]), scaling ([0.80, 1.20]), changing contrast, multiplying pixel values ([0.75, 1.25]), cropping ([0%, 2.5%] from all sides) and blurring (with probability 0.33 and blurring kernel [0.1%, 0.5%] of the image size). Second, the AAC scores for each augmented image were sampled from the approximated AAC6 distributions. The number of times an image was augmented and its AAC6 scores sampled depended on the mean AAC24 value of the image. The distribution of the mean AAC24 scores was very unbalanced and skewed towards the low AAC24 values, and hence the images with higher AAC24 values were augmented and their AAC6 scores sampled more often than images with low values to balance the disproportion in the training and validation sets (Fig. 4). Image augmentation was done for both training and validation set images, but the sampling of AAC6 outputs was done only for training set and for validation set the mean values of the AAC6 normal distributions were assigned as the outputs.

**Figure 4:**
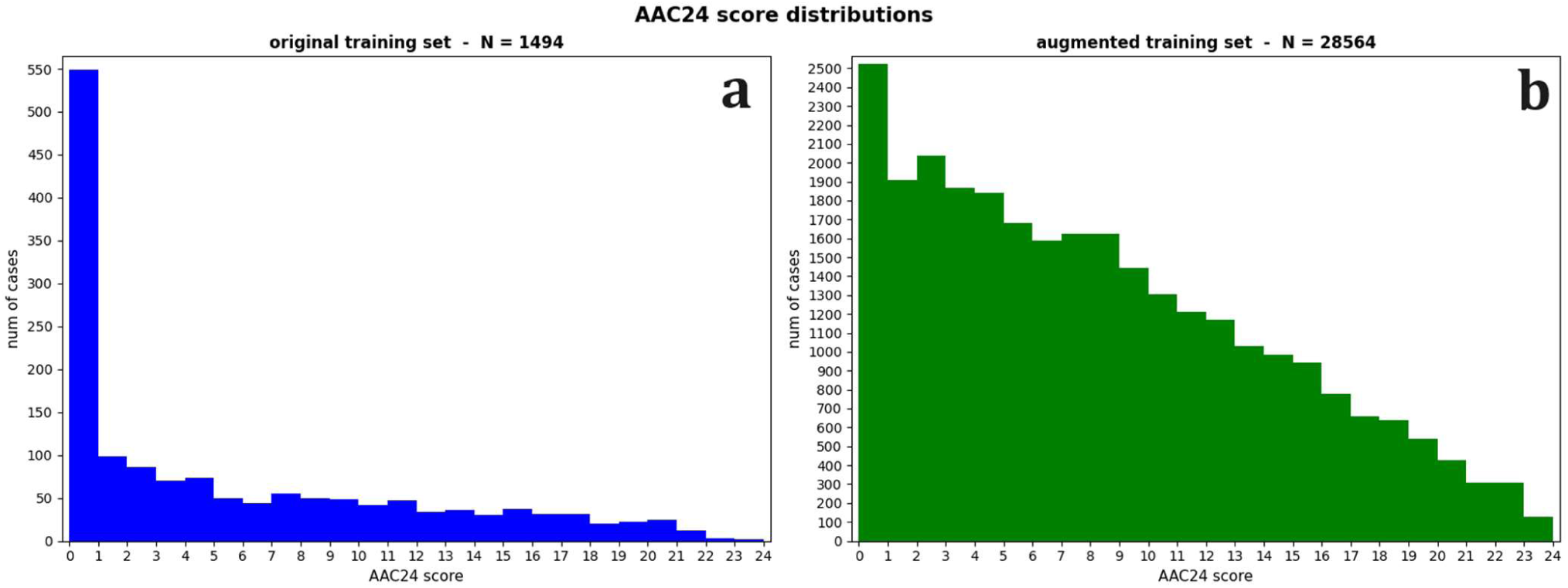
(a) The original and (b) augmented mean AAC24 distributions for one training set division (9 folds). Augmentation helped to reach more balanced distribution for AAC24 scores of the training and validation sets.

The architectural choices were based on performance on the internal test set. The selection process was done using X-ray images, and the same model architecture and settings were used with DXA images. Each model in the ensemble had the following architecture: a VGG-19 *[36]* type CNN part followed by a flattening layer and three fully connected (FC) layers with 128, 128 and 4 nodes. The final output FC layer corresponded to the output AAC6 scores. A sigmoid activation function was used in the output layer and rectified linear unit activation function in all the other layers. Dropout was used in the first two FC layers with a 0.30 dropout rate. Last, mean absolute error was used as the loss function.

In the training of the models, the VGG19-backbone was initialized with pre-trained ImageNet weights and all layers were trained. Adam optimizer was used as the optimization algorithm. A model was trained for 24 epochs using a batch size of 32, an initial learning rate of 0.75e-4 and the learning rate was reduced by 10% after every 4 epoch. The training was stopped if validation loss did not improve in 10 epochs. The models were built and trained using Python 3.9 and TensorFlow 2.7. Two RTX 6000 Ada GPUs with 48GB of display memory were used in the training.

The only difference between X-ray and DXA models was in the input images. DXA had three input channels: low energy, high energy and BMD images, whereas the X-ray had only one channel. The input image sizes were 448×448×1 for X-ray images and 320×320×3 for DXA images. For X-ray images, the chosen size was a compromise between resolution and the time required for model training. For DXA images, no downscaling was needed since the cropped images were not larger than 320×320.

### 2.5 Statistical tools for AAC score agreement analysis

To evaluate the variability and agreement between AAC scores made by radiologists or our ensemble model, we employed two statistical measures: the Concordance Correlation Coefficient (CCC) and the Total Deviance Index (TDI) (see, e.g., *[37, 38, 39]*). These two metrics provide a complementary interpretation of the repeatability of the AAC scoring process.

The CCC is a measure that combines accuracy and precision to evaluate the agreement between two continuous variables. CCC assesses how well pairs of (repeated) measurements are in concordance relative to each other. A CCC value of 1 indicates perfect concordance and agreement between the measurements, while a value of −1 indicates perfect discordance.

TDI quantifies the overall deviation between repeated measurements by focusing on the absolute differences between them. The TDI highlights the extent of variability and bias present in the measurements, providing a single measure for total error. The TDI is defined as the *p*-th percentile of the absolute differences between repeated observations. In this study, we chose *p* to be 90 (like in *[40]*). Hence, the interpretation of the index is: 90% of the absolute differences between the measurements fall within the calculated TDI value, offering a clear and interpretable metric of agreement. Lower TDI values indicate better agreement and less overall deviation between measurements.

In this study, we estimated CCC and TDI for intra- and inter-rater comparisons across the AAC score annotations but also for inter-rater comparison between annotations and model ensemble predictions by using a linear mixed model to take different dependencies adequately into account *[37]*.

## 3. Results

### 3.1 Agreement analysis for AAC annotations

Table 1 summarizes the CCC and TDI estimates for manual AAC scoring from X-ray images of the external test set within and between the three radiologists. Table 1 also reports the lower bound of 95% confidence interval (CI) for CCC and the upper bound of 95% CI for TDI. Three radiologists annotated each image twice, resulting in 6 annotations. Intra-annotator agreement and correlation are high for all three radiologists even though two of the radiologists outperformed the third: the CCC estimates range from 0.984 to 0.951 and the lower bounds of the 95% CIs were between 0.981 and 0.940. The 90% TDI estimates indicate a similar interpretation than CCC in the performance between the radiologists, estimates and the upper bounds of the 95% CIs ranging from 2.053 to 3.672 and 2.215 to 3.961, respectively. Even though the intra-annotator correlation is relatively high, the range that captures 90% of the differences in the annotations covers 9 to 16.5% of the AAC24 scoring scale (roughly 2.20 to 4.00).

**Table 1:** The intra- and inter-rater concordance correlation coefficients (CCCs) and total deviance indices (TDIs) for the AAC24 score annotated from X-ray images of the external test set for the three radiologists (RLs). Values are the estimates, and in the parenthesis are the lower bound of the 95 % confidence interval for CCC and upper bound of the 95 % confidence interval for TDI.

| AAC24 | CCC |  |  | TDI |  |  |
| --- | --- | --- | --- | --- | --- | --- |
| Annotator | RL1 | RL2 | RL3 | RL1 | RL2 | RL3 |
| RL1 | 0.984 (0.981) | 0.906 (0.881) | 0.895 (0.867) | 2.053 (2.215) | 5.067 (5.512) | 5.485 (5.944) |
| RL2 | - | 0.951 (0.940) | 0.900 (0.873) | - | 3.672 (3.961) | 5.286 (5.701) |
| RL3 | - | - | 0.978 (0.973) | - | - | 2.387 (2.575) |

All the inter-annotator CCCs between different radiologists are around 0.900 (Table 1). This shows a good correlation between the radiologists, even though the inter-annotator correlation is weaker than intra-annotator correlation. On the other hand, inter-annotator TDIs have a more than twice wider fluctuation range compared to intra-annotator values, showing lower agreement and higher variability. The range that includes 90% of the differences can cover almost 25% of the AAC24 scale, which is a relatively large range of variation. Additionally, mean absolute errors between the mean AAC24 scores of the three radiologists’ repeated annotations range from 1.704 to 2.423. The comparisons between the radiologists’ mean AAC24 scores are visualized in Supplementary material Fig. 2 (SMFig. 2).

The CCC and TDI estimates between the AAC24 scores of X-rays and DXA images are reported in Table 2. The repeated annotations by three radiologists were treated in this comparison between X-ray and DXA images as independent repeated measurements. The agreement and repeatability of the AAC score are better for X-ray images than for DXA images. The inter-rater agreement between X-ray and DXA annotations shows further reduction (CCC: 0.846 (0.822) and TDI: 6.639 (6.844)) compared to the agreement within a single modality. Clearly, as shown by the intra-rater CCC and TDI estimates, the AAC annotations are the most consistent when done from X-ray images, and the differences in the AAC scores annotated from X-ray and DXA can be relatively large in the AAC24 scale as indicated by the inter-rater TDI estimate.

**Table 2:** X-ray and DXA AAC annotation comparison: Intra- and inter-rater concordance correlation coefficients (CCCs) and total deviance indices (TDIs) for the AAC24 scores (6 scores in total per imaging modality) annotated from X-ray and DXA images of the external test set. Values are the estimates, and in the parenthesis are the lower bound of the 95 % confidence interval for CCC and upper bound of the 95 % confidence interval for TDI.

| AAC24 | CCC |  | TDI |  |
| --- | --- | --- | --- | --- |
| Image | X-ray | DXA | X-ray | DXA |
| X-ray | 0.904 (0.888) | 0.846 (0.822) | 5.165 (5.344) | 6.639 (6.844) |
| DXA | - | 0.853 (0.831) | - | 6.582 (6.808) |

### 3.2 AAC prediction with model ensemble

Fig. 5 (left) shows the predicted mean AAC24 values of the model ensemble for the X-ray and DXA images when compared with the mean annotated scores. The predicted scores from both modalities are in good agreement with the annotated scores: the scores predicted from X-ray have a mean absolute error of 1.058, and that from DXA increases to 1.491. The error distributions of the annotated and predicted mean AAC24 scores (Annotated*_mean_* -Predicted*_mean_*) presented in Fig 5. (right) shows that the mean error in both modalities is close to zero whereas the standard deviation of the error for predictions from DXA (2.069) is 30 % larger than that for X-ray (1.545).

**Figure 5:**
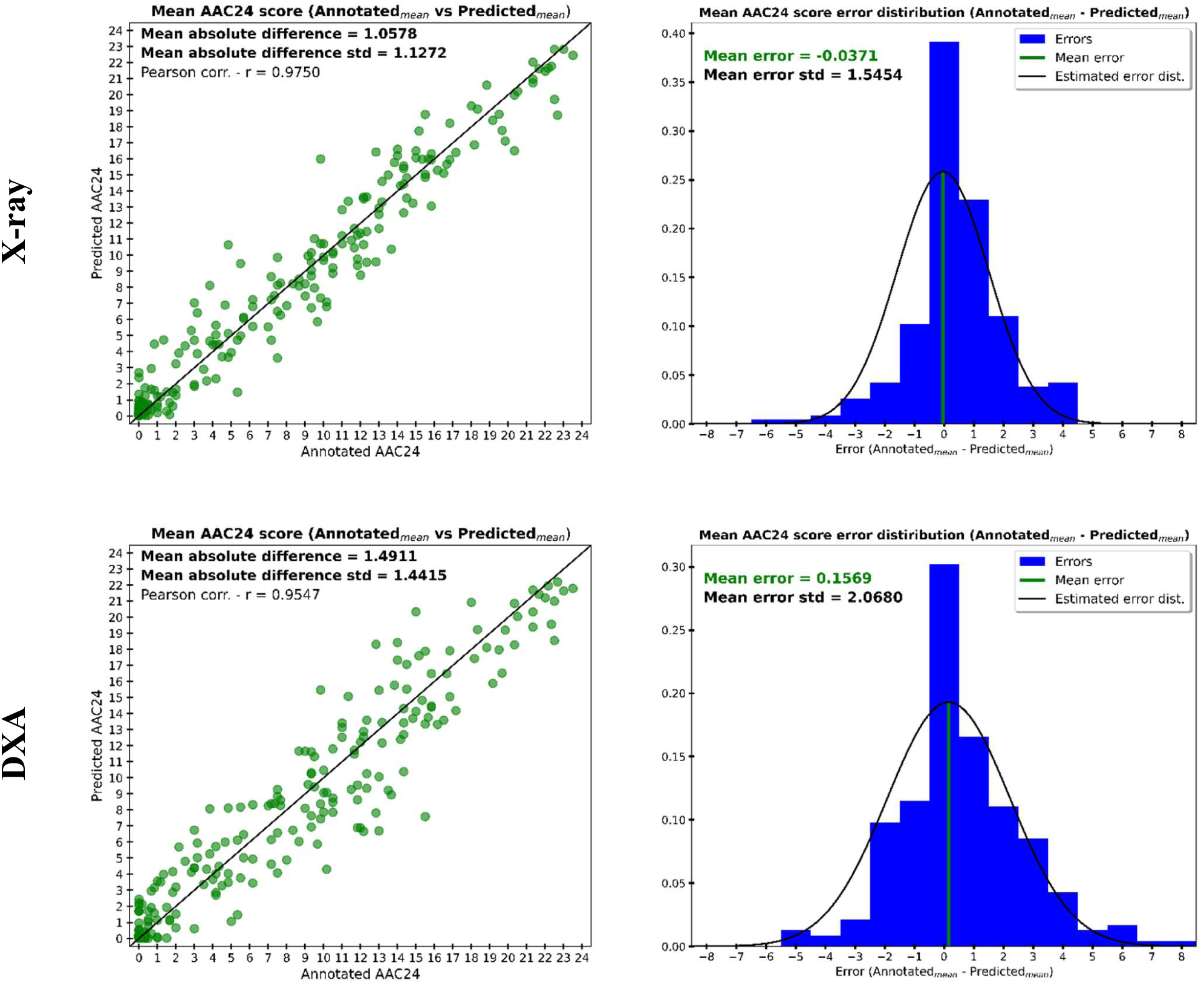
Left: The mean AAC24 scores for the annotations and ensemble model predictions for the X-ray (upper row) and DXA (lower row) images of the external test set. Right: The error distributions of the annotated and predicted mean AAC24 scores (Annotated_mean_ – Predicted_mean_). The mean error is close to zero for both modalities, but the standard deviation of the error is 30% higher for DXA than X-ray.

Fig 6. visualizes the 95% CIs of the annotation distributions and all the predicted AAC24 scores from the model ensemble for the external test set. The figure aims to give insight into how the annotations and ensemble predictions agree for X-ray and DXA images. There are a few outliers in the distribution estimates from both modalities as already seen in Fig 5. (left) but in most cases the ensemble predictions overlap with the 95% CIs of the annotations, and the average predictions and annotations show strong agreement. The variation in the AAC24 scores seems to be larger at the moderate AAC24 range when compared to low and severe AAC24 cases. Fig 6. also confirms the slightly larger difference from annotated scores when the AAC24 is predicted from DXA than X-ray. Nevertheless, the mean absolute error between the mean of annotated AAC24 scores and predicted from DXA (1.491) was smaller than the mean absolute errors between radiologists (1.704 to 2.423) and well within both the inter-rater (5.067 to 5.485) and intra-rater (2.053 to 3.672) TDI estimates reported in section 3.1.

**Figure 6:**
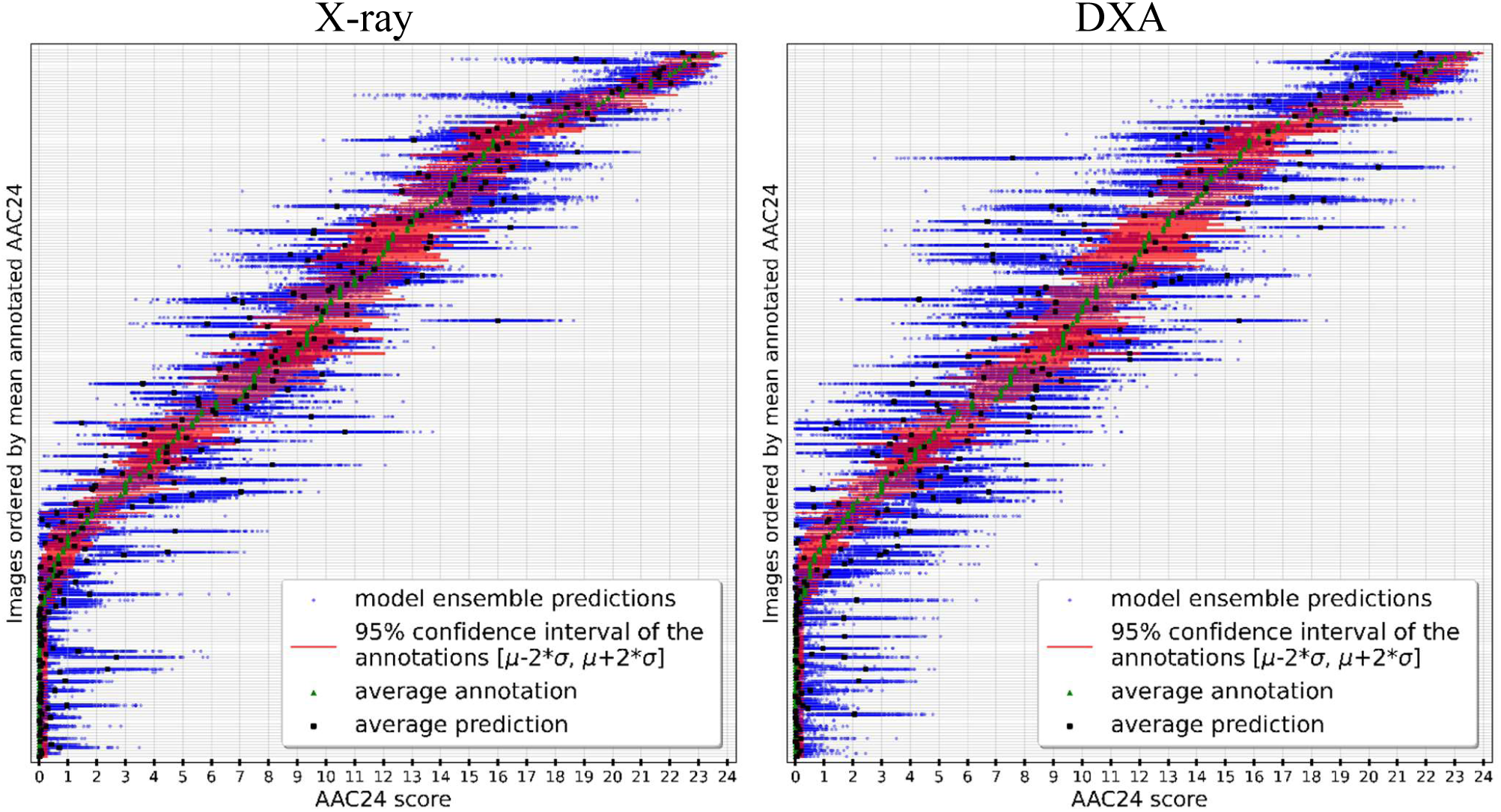
Left: X-ray and right: DXA. The 95% confidence intervals of the annotations and all the predicted AAC scores from the model ensemble for the external test set. The rows are ordered according to the mean annotated AAC24 score. Each row corresponds to one image and the horizontal axis ranges over the AAC24 score. Individual AAC24 predictions in the ensemble are presented with blue dots and their average with black square. 95% confidence interval of annotations are shown with red line and their average with green triangle.

Fig 7. shows typical examples of X-ray and DXA images with low, moderate, and high AAC. More example cases are provided in the Supplementary material. The annotated and predicted AAC distributions show good agreement for both imaging modalities in all AAC ranges.

**Figure 7:**
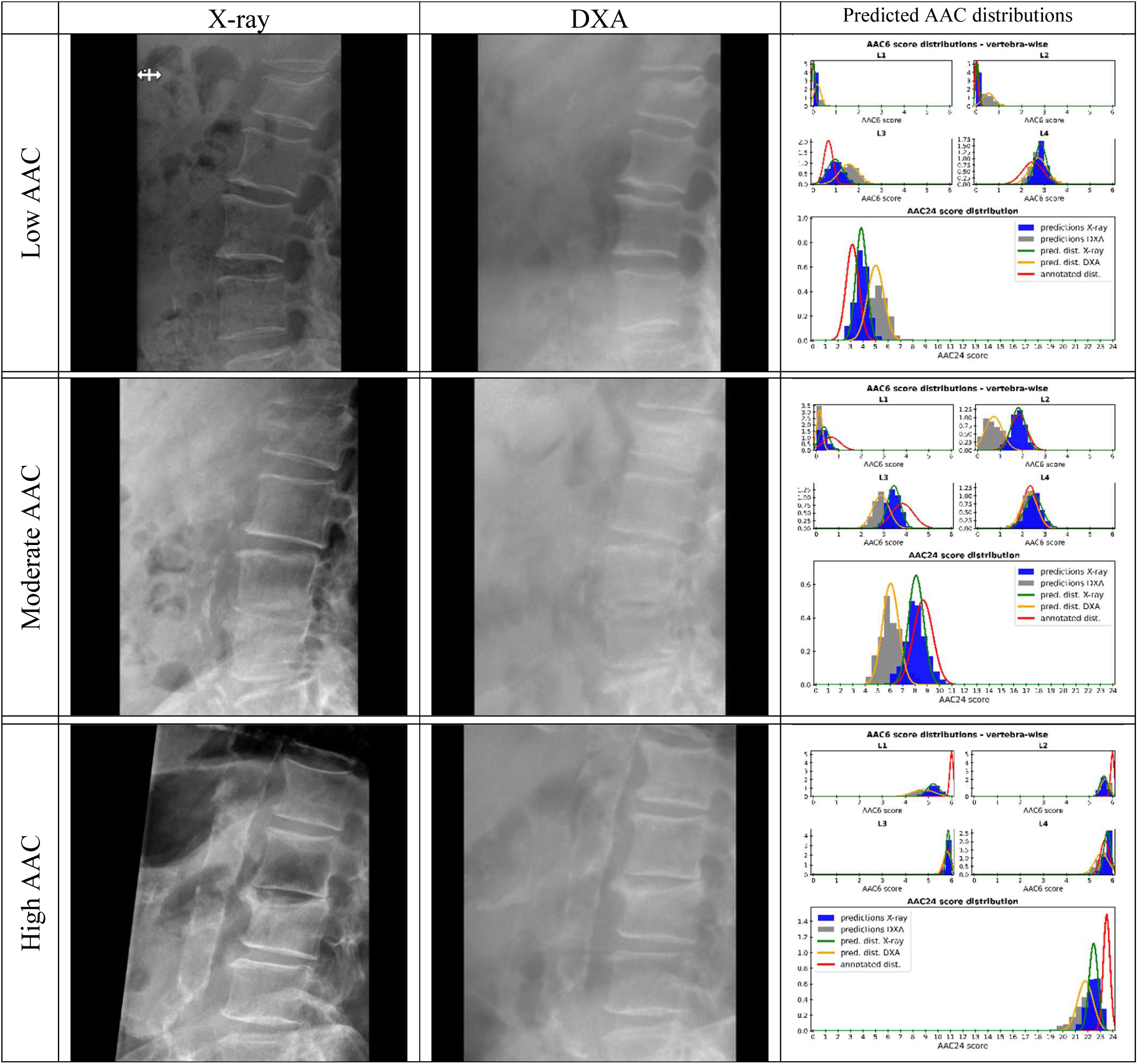
Left: Examples of cropped X-ray and DXA images with low, moderate and high AAC. Right: The annotated (red) and predicted (green for X-ray and yellow for DXA) AAC6 distributions and the resulting AAC24 distributions for both modalities.

Overall, the prediction accuracy drops only marginally for the external test set when compared to the internal test set. The internal test set results are presented in the Supplementary material. The mean absolute error between the annotated and predicted mean AAC24 scores increases from 0.864 to 1.058 for X-ray and from 1.350 to 1.491 for DXA. Also, the standard deviations of the mean AAC24 error increase from 1.409 to 1.545 for X-ray and from 2.024 to 2.068 for DXA, indicating a fraction larger deviation between the annotated and predicted values in the external test set.

We calculated CCC and TDI inter-rater agreement between all the radiologists’ AAC annotations and a random sample set from model ensemble predictions. In the agreement calculations, we included all AAC24 scores made by the three radiologists and took six random scores from the AAC24 ensemble predictions. The CCC (estimate (lower bound of 95% CI)) and TDI (estimate (upper bound of 95% CI)) between annotations and predictions were 0.930 (0.916) and 4.271 (4.459) for X-ray, respectively and for DXA 0.912 (0.895) and 4.841 (5.084). For both modalities, these CCC and TDI estimates showed better agreement than the inter-rater agreement among the individual radiologists reported in section 3.1.

### 3.3 Comparison of the proposed approach to previously published methods

Table 3 summarizes a comparison of correlation and error metrics for the predicted and annotated AAC scores at the L1-L4 vertebra levels between our approach, Gilani et al. *[28]* and Ilyas et al. *[30]*. They both used 1916 single and dual-energy DXA images to predict AAC scores at the L1-L4 vertebra-levels (AAC6) but do not report results for the cumulative AAC24. For our study, we summarize both X-ray and DXA image results at the vertebra levels and for the cumulative AAC24. The vertebra level AAC6 estimation results, presented as figures similar to AAC24 Fig. 5 in section 3.2, are shown in the Supplementary material. The correlation coefficients for DXA images in our approach outperformed those from the previous studies and the mean absolute errors were similar or lower on all vertebra levels. Our results on X-ray images are clearly the best based on reported metrics when compared to results on *[28]* and *[30]* but this may be explained by the different modalities used and that the manual AAC annotations in those works where done from DXA images.

**Table 3:** Summary of AAC estimation at the L1-L4 vertebra-levels of the proposed approach on X-ray and DXA images and methods *[28]* and *[30]* on DXA images. The arrows ↑ and ↓ indicate if higher or lower values are better for the corresponding metric.

| *Dual-Energy DXA Dataset - GE iDXA (1,916 images)* |  |  |  |  |  |  |  |
| --- | --- | --- | --- | --- | --- | --- | --- |
| **Evaluation metric** | **Method/Paper** | L1-AAC6 | L2-AAC6 | L3-AAC6 | L4-AAC6 | total | AAC24 |
| Pearson Correlation ↑ | Gilani et al. [28] | 0.49 | 0.64 | 0.70 | 0.69 | | |
|  | Ilyas et al. [30] | 0.77 | 0.80 | 0.83 | 0.84 |  |  |
| Mean absolute error ↓ | Gilani et al. [28] | 0.60 | 0.67 | 0.88 | 1.10 | 3.25 | |
|  | Ilyas et al. [30] | 0.43 | 0.47 | 0.69 | 0.70 | 2.29 |  |
| *Single-Energy DXA Dataset - GE iDXA (1,916 images)* |  |  |  |  |  |  |  |
| Pearson Correlation ↑ | Ilyas et al. [30] | 0.79 | 0.81 | 0.84 | 0.85 | | |
| Mean absolute error ↓ | Ilyas et al. [30] | 0.40 | 0.45 | 0.67 | 0.67 | 2.19 | |
| *Kuopio external test set - X-ray/iDXA (235 images)* |  |  |  |  |  |  |  |
| Pearson Correlation ↑ | X-ray ensemble mean | 0.92 | 0.95 | 0.95 | 0.96 | | 0.97 |
|  | DXA ensemble mean | 0.86 | 0.92 | 0.93 | 0.91 |  | 0.95 |
| Mean absolute error ↓ | X-ray ensemble mean | 0.33 | 0.36 | 0.45 | 0.44 | 1.58 | 1.06 |
|  | DXA ensemble mean | 0.43 | 0.46 | 0.55 | 0.65 | 2.09 | 1.49 |

## 4 Discussion

Abdominal aortic calcification is a surrogate for future cardiovascular events, but its scoring systems hold inherent variation. We therefore addressed the uncertain nature of individual AAC scores with an ensemble learning approach, where models are trained with scores sampled from a distribution of probable scores derived from manual annotations. The approach adds to the previously reported automated AAC scoring methods on X-ray *[22, 23]* and DXA *[24, 25, 26, 27, 28, 29, 30, 31, 32]*, since to the best of our knowledge all previous methods treat the AAC score as a single deterministic value. This has been so even though according to literature radiologists do not always agree when scoring findings from projection images *[41, 40]*.

Our analysis of the manually annotated AAC scores confirmed, similarly as has been reported, e.g., in *[16]*, that there exists differences when radiologists annotate AAC scores using the widely adopted 24-point scoring system *[11]*. Agreement analysis presented in Table 1 showed that the variability and interpretation differences between the scorers can cover up to 25% of the 24-point scale, highlighting the need not rely solely on single scores.

The proposed approach to take the variability in the AAC scores into account proved beneficial, since the predicted AAC distributions showed good agreement with the consensus scores of the radiologists. Importantly, the agreement was better than the inter-rater agreement between individual radiologists (cf. Table 1 and section 3.2 and Fig. 5 vs. SMFig 2), which can be seen as a reference level of accuracy. These results indicate that the proposed ensemble approach allows us to predict AAC scores with higher accuracy level than how two radiologists perform in the scoring task relative to each other.

The automatic scoring in this work yielded better accuracy metrics than reported for similar methods in the literature. The comparison presented in section 3.3 showed that evaluation metrics (correlation and error) were better or similar for our method on the external test set than the authors of previous methods had reported for their test sets. It needs to be noted that the evaluation metrics of the X-ray based method were not fully comparable to the other methods designed for DXA based scoring. However, we argue that for DXA-based method the comparison was fair although datasets were different, since the images were obtained with the same scanner type (GE Lunar iDXA). This comparison further supported the benefits of our approach to treat the scored AAC as a normally distributed random variable and not as a single deterministic value.

AAC scores were annotated from X-ray images, because we had no access to a large enough dataset with CT, X-ray and DXA image triplets that would have had scan dates within 365 days, which was used as the inclusion criterion. AAC scores annotated from X-ray and DXA images can have a relatively large difference (90% TDI = 6.639 (Table 2.)), and therefore the scores would not be directly comparable. DXA images may miss smaller amounts of calcification that are still detected by X-ray due to the use of typically one-tenth of radiation dose and lower resolution compared to X-ray imaging. Thus, due to better sensitivity, AAC scored from X-ray images was considered the underlying ground truth when training AAC predictions models for both X-ray and DXA images. This approach allowed for a direct comparison of results between the two modalities, which would have been inconvenient if the ground true scores were obtained separately for X-ray and DXA. The accuracy of AAC prediction was overall slightly better from X-ray images than from DXA. This, however, was expected due to the lower resolution in DXA imaging and that the ground truth AAC scores were annotated from X-ray.

The X-rays in the external test set were imaged with a variety of X-ray devices from different manufacturers, some of which were not represented in the training set. The results suggest that the AAC estimation model generalizes well to X-rays imaged with different devices. However, the DXA results have a limitation that the ensemble was trained and tested with images from devices of a single manufacturer. Nevertheless, based on the good performance on multi-device X-ray data, the prediction accuracy for DXA images from multiple vendors is not expected to drop considerably. Another indication of generalization capabilities of our AAC estimation approach, is that for external test set the prediction error between the annotated and predicted mean AAC24 values increased only about 10-15% compared to internal test set.

Time requirement to establish and train a model ensemble, is indeed considerable. The approach requires multiple manual AAC annotations to approximate the AAC statistics. Training an ensemble of 400 models takes multiple weeks where a single CNN regression model can be trained on a desktop computer within 1-1.5 hours. However, when the model ensemble is trained, the inference time to predict a new AAC score distribution estimate can be done in seconds, if single model predictions, which are independent of each other, are parallelized effectively.

The results show that the mean value of the predicted ensemble scores is a good point estimate for the AAC. Additionally, since the model ensemble outputs a distribution of AAC predictions, one can also estimate a CI for the point estimate. For example, since the ensemble predictions are assumed to approximate a normal distribution, one can further approximate CI based on the mean and standard deviation of the ensemble predictions. Another feasible choice to construct and approximate CIs could be, for instance, bootstrap-type approaches. The width of the CI can be interpreted as a measure of uncertainty in the predicted AAC scores, which would be a useful addition for clinical applications of the method.

## 5 Conclusions

In this paper, we showed that an ensemble of convolutional neural network models can predict the amount of calcification in the abdominal aorta from lateral lumbar spine X-ray and DXA images at the lumbar vertebra L1 to L4 levels. Our approach was based on the idea that the abdominal aortic calcification (AAC) score is treated as a normally distributed random variable instead of just one point value and a model ensemble was trained to produce an ensemble of predictions to approximate the AAC distribution.

Overall, the results of this study demonstrate that deep learning-based AAC ensemble estimation from lateral lumbar spine X-ray and DXA images can be done with compatible accuracy and have strong correlation to manual annotations by radiologists. Moreover, the automated AAC estimation produces a distribution of possible AAC scores at the vertebra L1 to L4 levels from which point estimates, such as the mean, can be obtained, and further, confidence intervals (CIs) can also be estimated to describe the uncertainty. The proposed method to predict AAC could offer an opportunistic screening tool for multiple imaging modalities.

## CRediT author statement

**Antti Voss:** Writing - Original Draft, Writing - Review & Editing, Funding acquisition, Visualization, Conceptualization, Methodology, Software, Formal analysis, Investigation **Sanna Suoranta:** Methodology, Conceptualization, Investigation **Tomi Nissinen:** Writing - Original Draft, Writing - Review & Editing, Conceptualization, Investigation **Ossi Hurskainen:** Investigation **Masarwah Amro:** Investigation **Reijo Sund:** Writing - Review & Editing, Conceptualization, Formal analysis **Jussi Tohka:** Writing - Review & Editing, Conceptualization, **Sami P Väänänen:** Writing - Original Draft, Writing - Review & Editing, Supervision, Funding acquisition, Methodology, Conceptualization, Investigation

## Acknowledgements

We acknowledge the Research Committee of the Wellbeing services county of North Savo for the State Research Funding (projects 5203101 and 5063582). This study was also supported by Research Council of Finland (projects 2021/ 338647, 2023/ 356743 and 358944 (Flagship of Advanced Mathematics for Sensing, Imaging and Modeling)).

## Declaration of competing interest

The authors declare that they have no known competing financial interests or personal relationships that could have appeared to influence the work reported in this paper.

## Data availability

The authors do not have permission to share data.

## Ethics statement

The pseudo anonymized patient data for the study was collected and used according to a research permission granted by the Finnish Institute for Health and Welfare and ethical permission granted by Research Ethics Committee of Hospital District of Northern Savo.

## Notes

### Competing Interest Statement

The authors have declared no competing interest.

### Funding Statement

This study was funded by the Research Committee of the Wellbeing services county of North Savo for the State Research Funding (projects 5203101 and 5063582) and the Research Council of Finland (projects 2021/ 338647, 2023/ 356743 and 358944 (Flagship of Advanced Mathematics for Sensing, Imaging and Modeling)).

### Author Declarations

Research Ethics Committee of Hospital District of Northern Savo gave ethical approval (date: 10.9.2019, reference number: 1298/13.02.00/2019) and the Finnish Institute for Health and Welfare gave permission (date: 18.3.2020, reference number: THL/2247/5.05.00/2019) to collect and use pseudo anonymized patient data (images, other patient information) in this work.

