## Supplementary material for "Deep learning ensemble for abdominal aortic calcification scoring from lumbar spine X-ray and DXA images"

This is the supplementary material of the article "*Deep learning ensemble for abdominal aortic calcification scoring from lumbar spine X-ray and DXA images*".

The supplementary material includes four sections: The first section presents how the aortic region was located and cropped from the original X-ray and DXA images based on estimated landmark points. The second section presents scatter plots between the mean AAC24 scores of the three radiologists' repeated manual annotations. Section 3 shows the main results of AAC prediction with model ensemble for the internal test set. Section 4 presents additional example cases of the AAC prediction on the external test set images. Section 4 also shows the vertebra level AAC estimation results for the external test set.

### 1. Materials and methods

#### Extracting the aortic region based on vertebra landmarks

The aortic region from X-ray and DXA images was located and cropped based on landmark points or levels that were estimated using CNN-based deep-learning models (see Fig. 1). For X-ray images, there were a total of 24 landmark points (6 points per vertebra) and a binary output (left/right) for which direction was the anterior side of the lateral image pointing. For DXA images, three levels were estimated: two y-coordinates for the superior part of L1 vertebra and the inferior part of L4 vertebra, and x-coordinate for the most posterior part of vertebrae L1 to L4. In DXA, the anterior side was always on the left. The low energy images were used to estimate landmarks from DXA images since anatomical structures are the clearest in the low energy images.

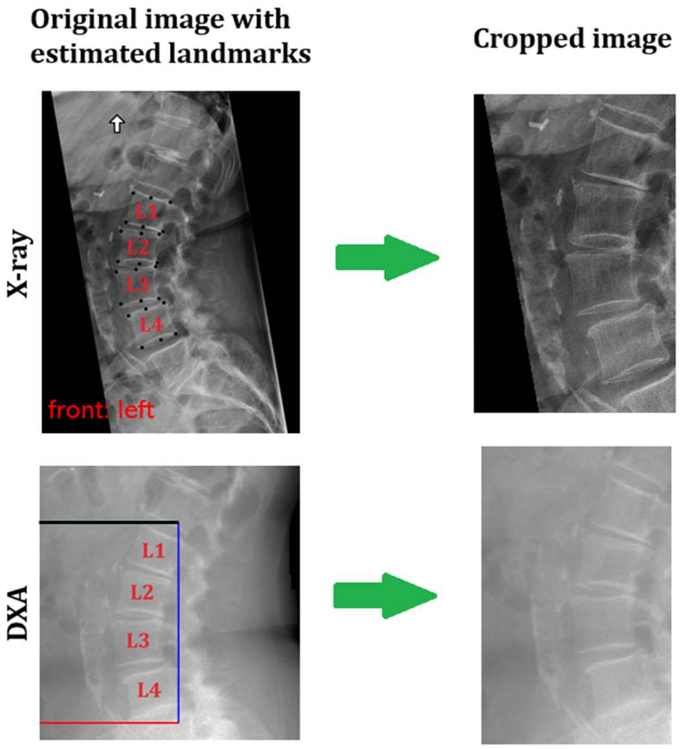


**Figure 1: Estimated landmarks and the corresponding cropped images for X-ray (top) and DXA (bottom) images. For X-ray images, there are 24 landmark points (black dots, 6 points per vertebra) and a binary output (left/right) indicating the anterior side of the lateral image. For DXA images, there are three landmark levels: two y-coordinates for the superior part of L1 vertebra (black line) and the inferior part of L4 vertebra (red line), and x-coordinate for the most posterior part of vertebrae L1 to L4 (blue line).**

The original images were cropped based on the estimated landmark points/levels such that vertebrae L1-L4 and the assumed aortic area were included. The X-ray images facing the right were first flipped horizontally. Then, the minimum and maximum values of the *x*- and *y*-coordinates of those estimated

24 points were taken. To get the final cropped area for an X-ray, the box was grown by 400 and 35 pixels to the anterior and posterior directions, and 120 pixels to the superior and inferior directions. The DXA images were cropped as follows: from the estimated *x*-coordinate we took 180 and 15 pixels to the anterior and posterior directions. From the estimated L1 superior and L4 inferior *y*-coordinates we took 30 pixels to the superior and inferior directions.

To train the landmark estimation models, we collected unpaired sets of 1948 DXA and 3308 X-ray images for training and 150 DXA and 299 X-ray images for internal testing among the patients from Joensuu and marked the landmarks on the images as described in this section. These images partly overlapped with the ones used to train the AAC prediction models, but the final performance evaluation of the proposed AAC estimation pipeline is done for the external test set from Kuopio where all the images are taken from a completely different hospital than the images that were used to train any of the models.

The architectures of the landmark estimation models for X-ray and DXA images were essentially the same, only varying in the number of nodes in the fully connected (FC) layers. The selections of the model architectures and parameters were based on the models' performance on the landmark internal test set. A model architecture that had the smallest mean absolute difference in landmark points/levels between the estimates and true landmarks was chosen.

The following model architectures were chosen: In both X-ray and DXA models a VGG19 convolutional neural network (CNN) was used for feature extraction. The X-ray model had a flattening layer and three FC layers with 1024, 1024 and 49 nodes after the CNN part as a regression part. The X-ray output layer included 48 normalized x- and y-coordinates for the vertebra landmark points and a binary value for the anterior side of the image (left/right – 0/1). The DXA model had a global average pooling layer after the CNN part and three FC layers with 256, 256 and 3 nodes as a regression part. The DXA output layer had two normalized y-coordinates for L1 and L4 vertebra levels and a normalized x-coordinate for the most posterior part of the L1-L4 vertebrae. In both models, sigmoid activation function was used in the output layers and rectified linear unit activation function in the other FC layers and in the CNN part. Mean absolute error was used as a loss function. Input image sizes of 320x320x1 and 288x288x1 were used for the X-ray and DXA models, respectively. To maintain the aspect ratio of the original images, they were first made square images by adding zeros and then resized to the desired size.

In the training process of both X-ray and DXA models, the VGG19-backbone CNN part was initialized with the pre-trained ImageNet weights and all layers were trained. Adam optimizer was used in the optimization process. To increase the size of the training data, image augmentation was applied and the following augmentation procedures were used: rotation ([-20°, 20°]), shear ([-7°, 7°]), translation (*x*:[-25%, 25%], *y*:[-20%, 20%]), horizontal flip (with probability 0.50 and X-ray only), scaling ([0.75, 1.25]), changing contrast, multiplying pixel values ([0.75, 1.25]), cropping ([0%, 5%] from all sides) and blurring (with probability 0.33 and blurring kernel [1%, 3%] of the image size). If any of the landmarks ended up outside the original image boundaries due to the affine transformations, the augmented image was discarded, and the augmentation was done again. Each X-ray and DXA images were augmented 10 times.

For both X-ray and DXA models, the training data was divided to 7 folds. For each set of fold divisions (6 folds for training and 1 for validation) we trained 4 models by re-doing the augmentation process every time. This resulted in a 28-model ensemble and the final landmark estimates were computed as median values of the model outputs. Each model was trained for 100 epochs using a batch size of 32, an initial learning rate of 1e-4 and the learning rate was reduced by 10% after every

10 epochs. The training was stopped if validation loss did not improve in 30 epochs.

### 2. Results: Agreement analysis for AAC annotations

Fig. 2 shows scatter plots between the mean AAC24 scores of the three radiologists' repeated annotations. The mean absolute errors between the radiologists' AAC24 scores range from 1.704 to 2.423.

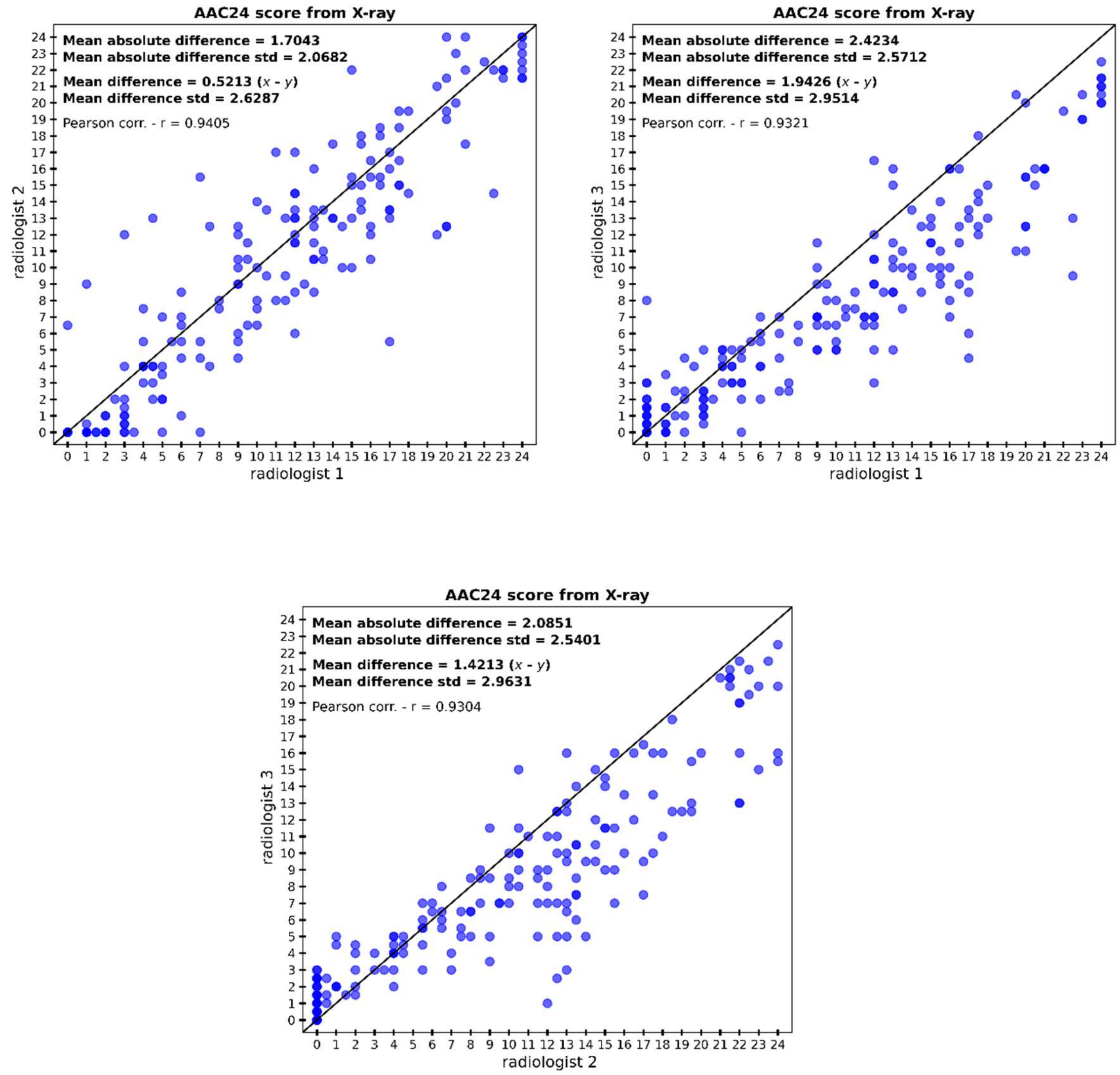


**Figure 2: External test set – Scatter plots of the annotated mean AAC24 scores between different radiologists (RLs): RL1 vs. RL2, RL1 vs. RL3 and RL2 vs. RL3. Mean absolute and mean difference and the corresponding standard deviations are given in each plot.**

### 3. Results: Internal test set

This section presents the results of the AAC estimation for X-ray and DXA images of the internal test set. The interpretation of the results is mostly based on the mean AAC24 values of the annotations and model ensemble predictions. In addition, we show a couple of low, moderate and high AAC example images and the corresponding predicted AAC distributions.

Fig. 3. (left) shows the mean AAC24 scores of the model ensemble predictions and the corresponding annotated mean scores for the X-ray and DXA images. There is a good agreement between the predicted and annotated mean AAC24 scores from both modalities: X-ray predictions have a mean absolute difference of 0.896 and for DXA predictions the difference is 1.350. The error distributions of the annotated and predicted mean AAC24 scores ($\text{Annotated}_{mean} - \text{Predicted}_{mean}$) presented in Fig 3. (right) show that the standard deviation of the error for DXA estimates (2.024) is about 40 % larger than for X-ray (1.409). This means larger variation in the AAC estimates from the annotated scores when the prediction is done from a DXA image.

Fig 4. visualizes the 95 % confidence intervals of the annotation distributions and all the predicted AAC scores from the model ensemble for X-ray and DXA images. Each row corresponds to an individual image (rows are ordered according to the mean annotated AAC24 score) and the horizontal axis indicates the AAC24 score. Fig 4. roughly illustrates how the annotations and ensemble predictions agree. There are a few outliers in the distribution estimates but in most cases the ensemble predictions overlap with the 95% confidence intervals of annotations, and furthermore, the average predictions and annotations have a good agreement.

Fig 5. shows examples of low, moderate and high AAC X-ray and DXA images and the resulting AAC ensemble predictions. In general, the annotated and predicted AAC distributions are in good agreement in both X-ray and DXA images in all the shown example cases.

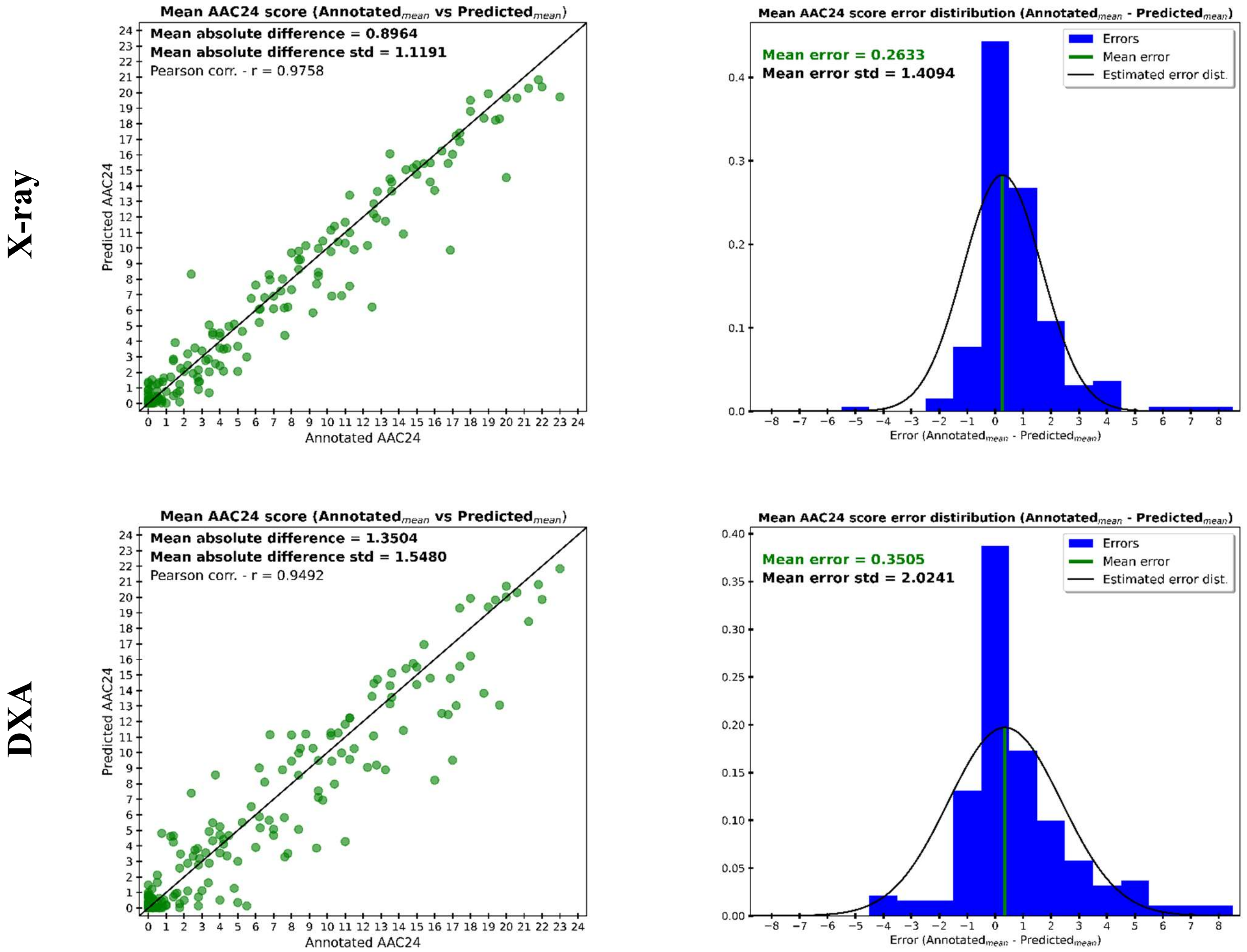


**Figure 3: Left: The mean AAC24 scores for the annotations and ensemble model predictions for the X-ray (upper row) and DXA (lower row) images of the internal test set. Right: The error distributions of the annotated and predicted mean AAC24 scores (Annotated$_{mean}$ – Predicted$_{mean}$).**

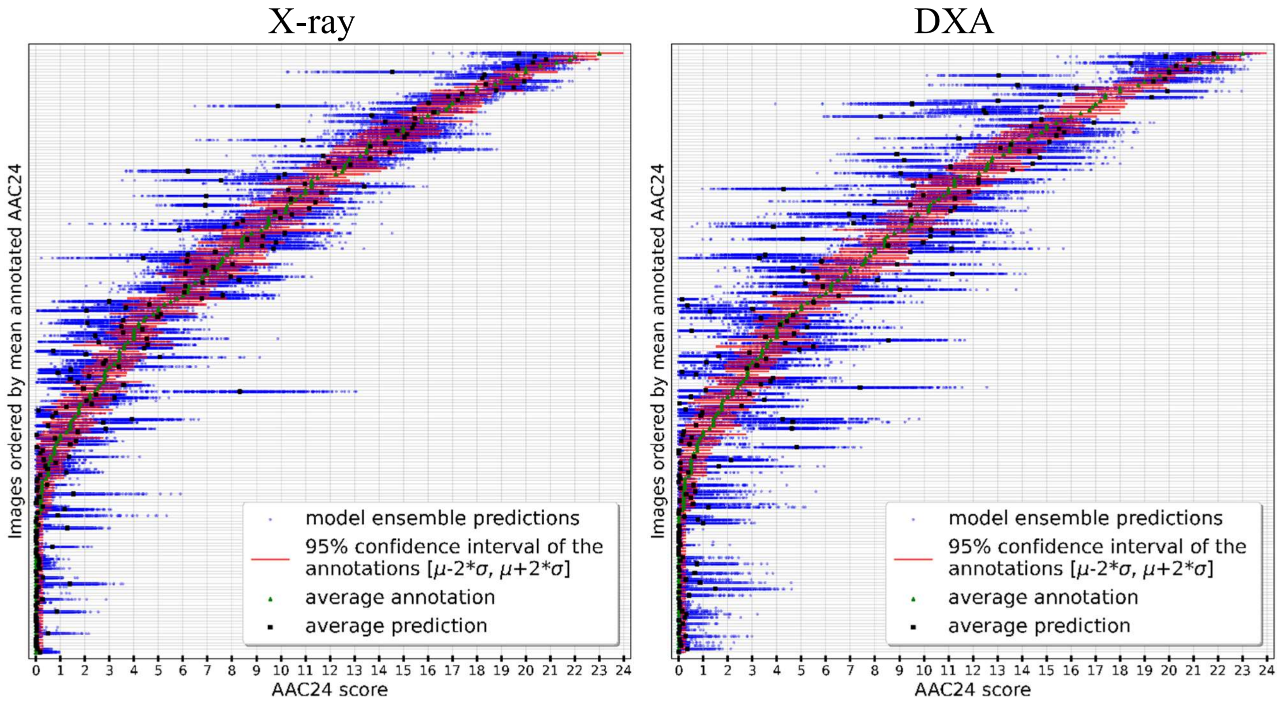


**Figure 4: Left: X-ray and right: DXA. The 95% confidence intervals of the annotations and all the predicted AAC scores from the model ensemble for the internal test set. The rows are ordered according to the mean annotated AAC24 score. Each row corresponds to one image and the horizontal axis ranges over the AAC24 score. Individual AAC24 predictions in the ensemble are presented with blue dots and their average with black square. 95% confidence interval of annotations are shown with red line and their average with green triangle.**

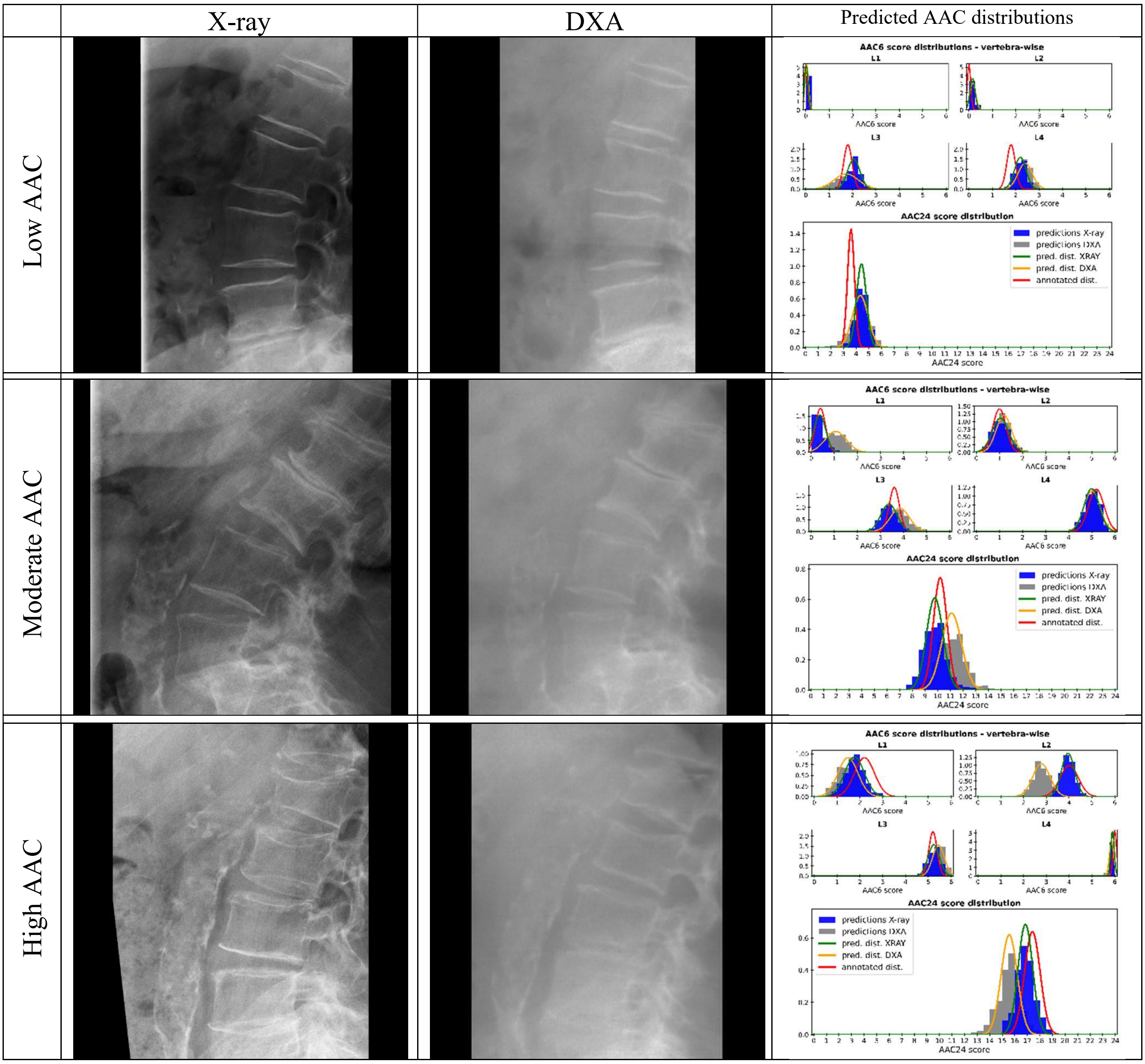


**Figure 5: Internal test set - Left: Examples of cropped X-ray and DXA images with low, moderate and high AAC. Right: The annotated (red) and predicted (green for X-ray and yellow for DXA) AAC6 distributions and the resulting AAC24 distributions for both modalities.**

### 4. Results: External test set – Additional example cases and vertebra level AAC predictions

In Figs 6.-8., 9 additional examples of low, moderate and high AAC X-ray and DXA images and the resulting AAC ensemble predictions are presented from the external test set. The L1-L4 vertebra level AAC estimation results are shown in Figs. 9 and 10. Fig 9. shows the vertebra-wise predicted mean AAC6 values of the model ensemble for X-ray and DXA images when compared with the annotated mean scores. The vertebra-wise error distributions of the annotated and predicted mean AAC6 scores ($Annotated_{mean}$ - $Predicted_{mean}$) are presented in Fig 10.

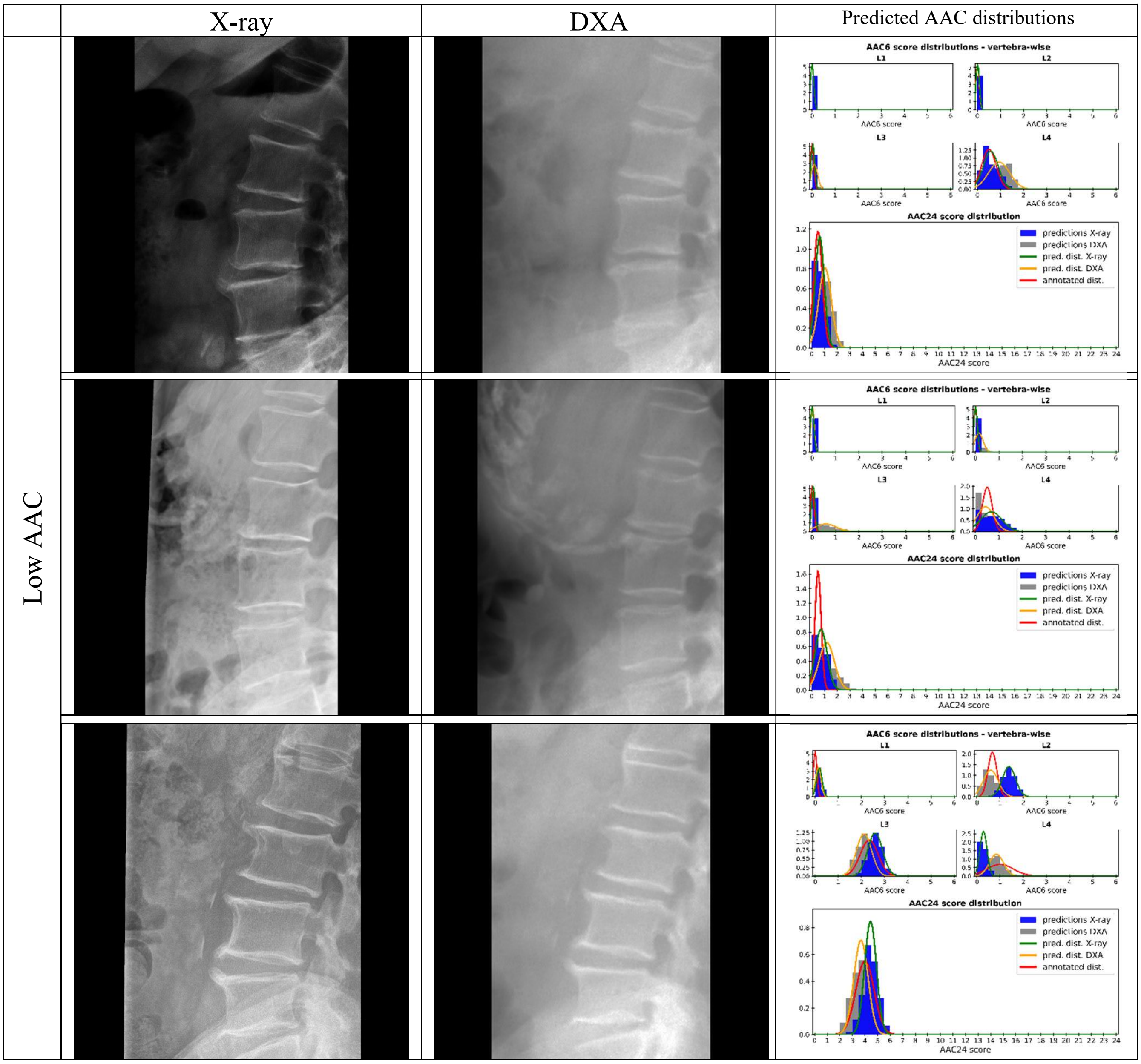


**Figure 6: External test set. Left: Examples of cropped X-ray and DXA images with low AAC. Right: The annotated (red) and predicted (green for X-ray and yellow for DXA) AAC6 distributions and the resulting AAC24 distributions for both modalities.**

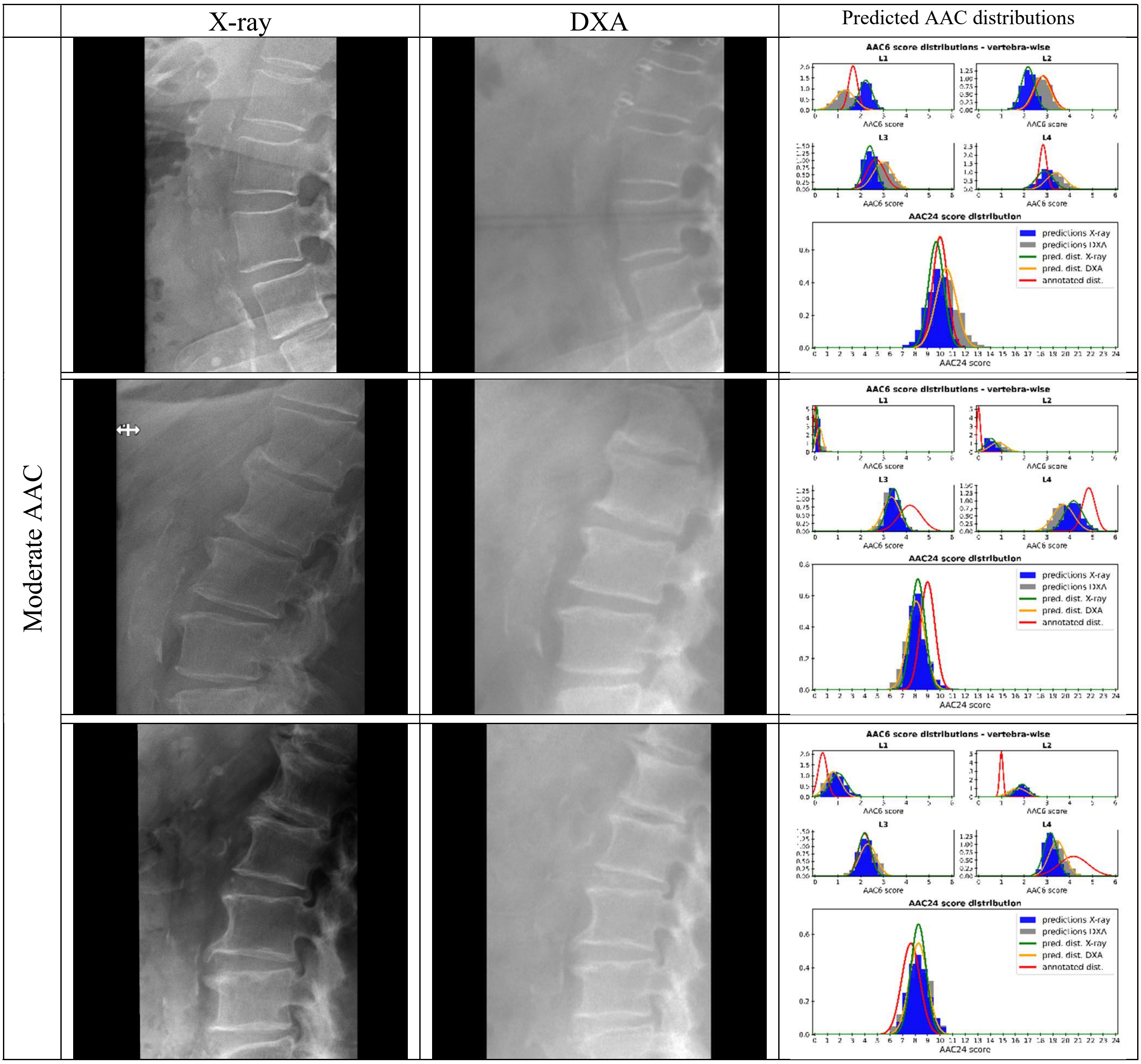


**Figure 7: External test set. Left: Examples of cropped X-ray and DXA images with moderate AAC. Right: The annotated (red) and predicted (green for X-ray and yellow for DXA) AAC6 distributions and the resulting AAC24 distributions for both modalities.**

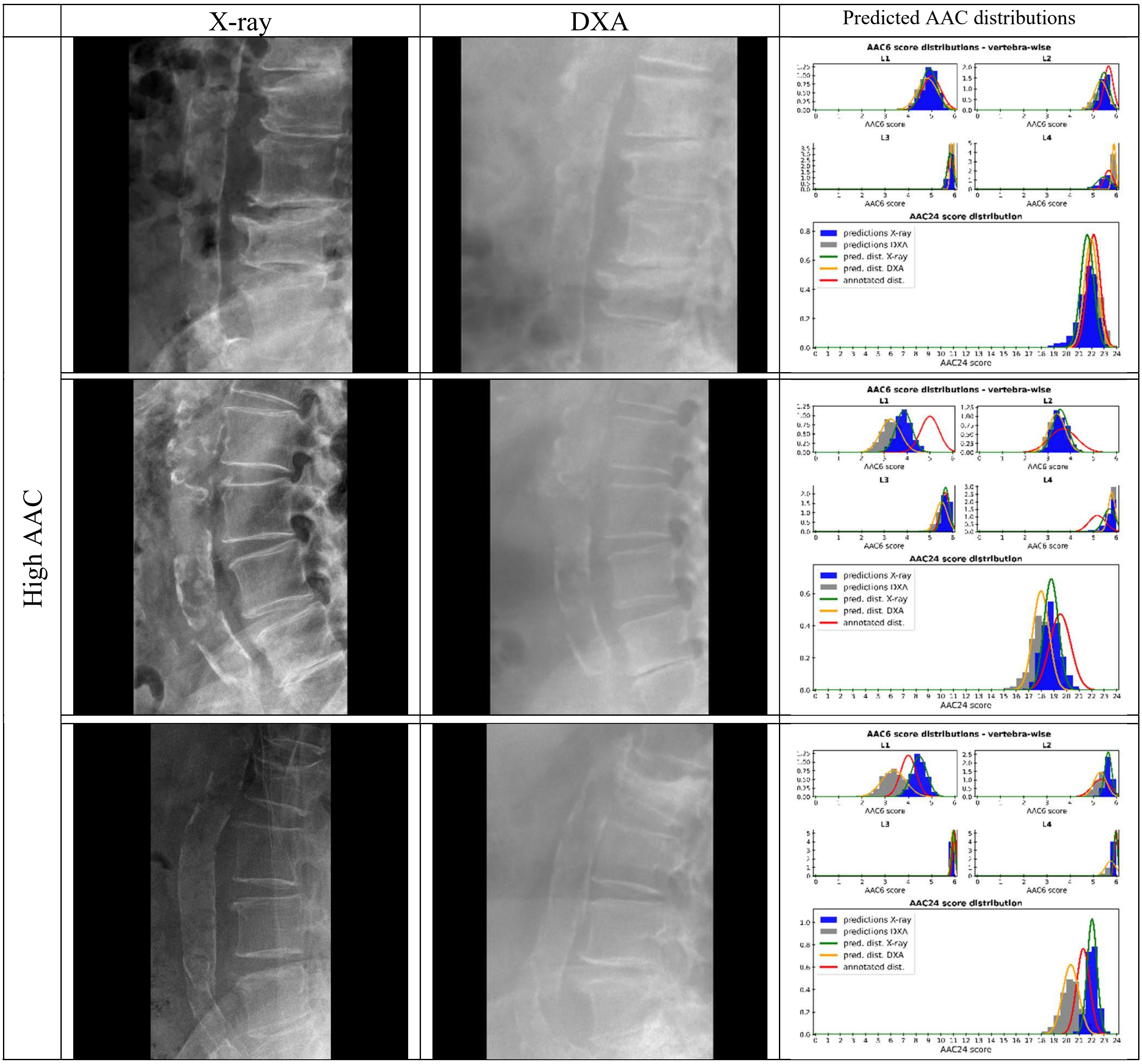


**Figure 8: External test set. Left: Examples of cropped X-ray and DXA images with high AAC. Right: The annotated (red) and predicted (green for X-ray and yellow for DXA) AAC6 distributions and the resulting AAC24 distributions for both modalities.**

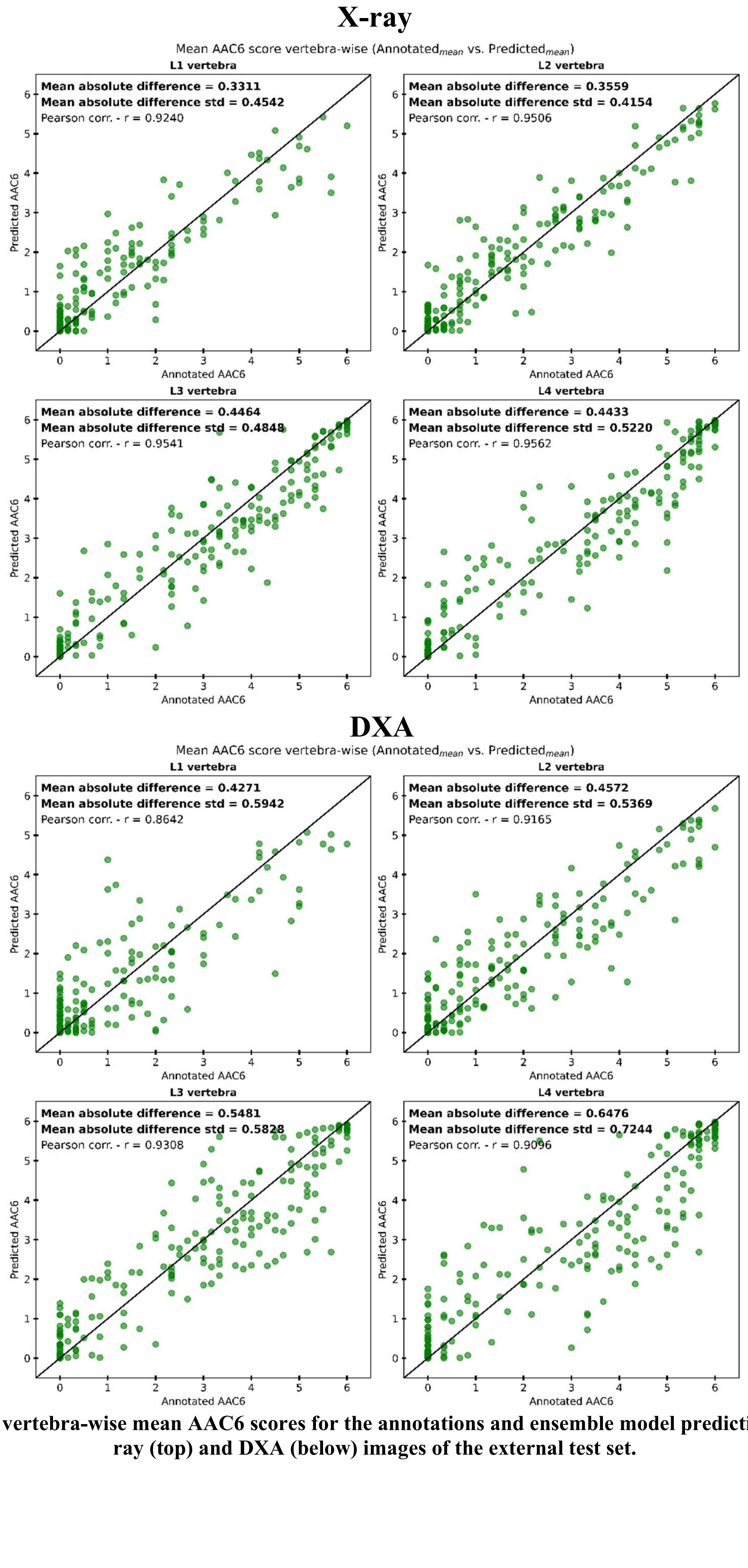


**Figure 9: The vertebra-wise mean AAC6 scores for the annotations and ensemble model predictions for the X-ray (top) and DXA (below) images of the external test set.**

### X-ray

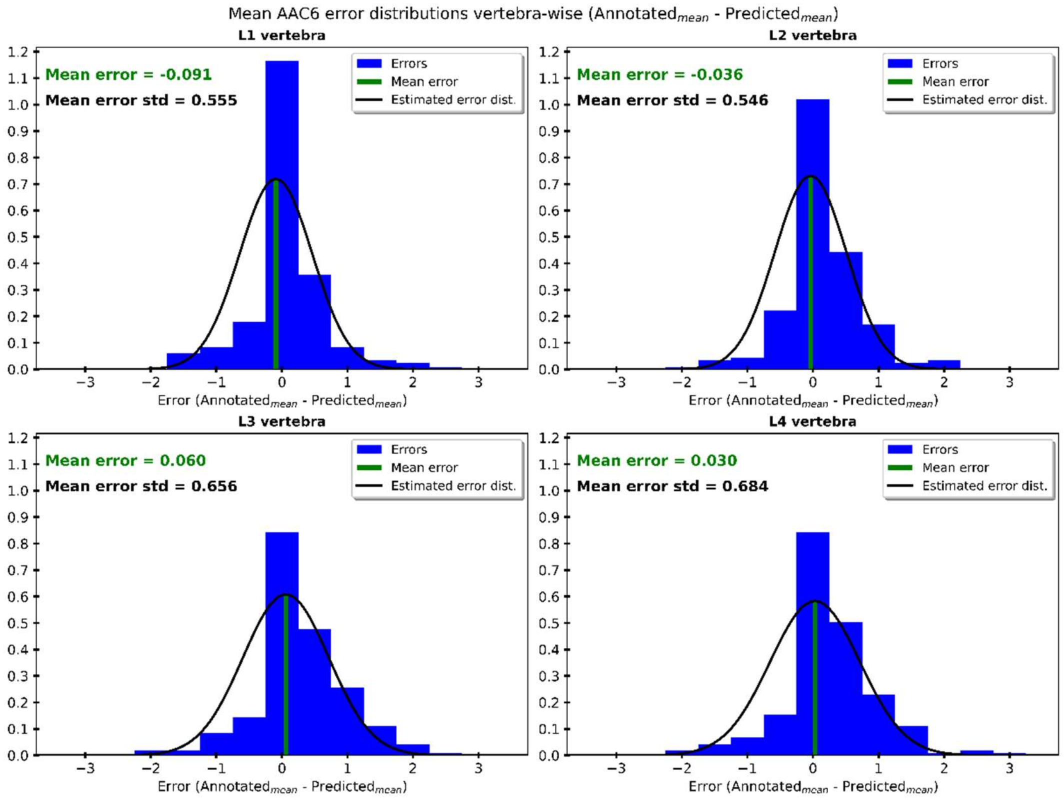


### DXA

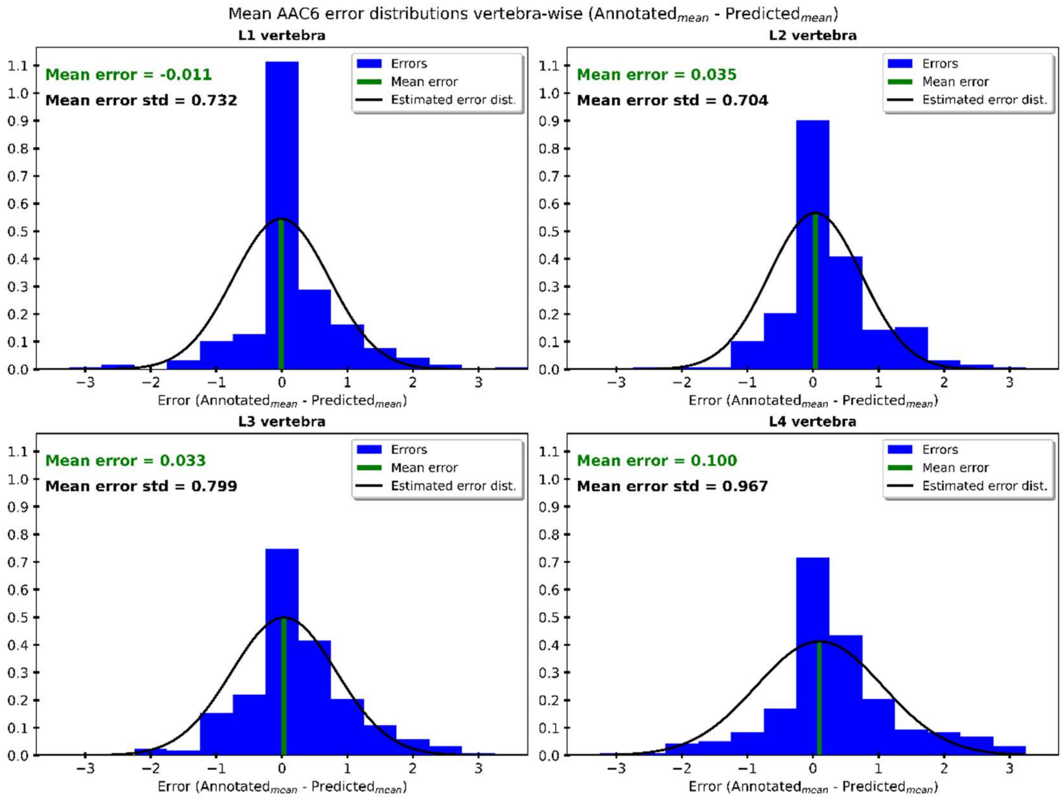


**Figure 10: The vertebra-wise error distributions of the annotated and predicted mean AAC6 scores (Annotated$_{mean}$ – Predicted$_{mean}$) for the X-ray (top) and DXA (below) images of the external test set.**
